# Evaluating the Association and Predictability of Complex Medication Regimen Scores with Clinical Outcomes Among the Critically Ill

**DOI:** 10.1101/2022.02.03.22270376

**Authors:** Mohamamd Al-Mamun, Jacob Strock, Yushuf Sharker, Rebecca Schmidt, Khaled Shawwa, Douglas Slain, Ankit Sakhuja, Todd Brothers

**Affiliations:** University of West Virginia, School of Pharmacy, Morgantown, West Virginia, United States of America; University of Rhode Island, Graduate School of Oceanography, Kingston, Rhode Island, United States of America; Children’s National Hospital, Washington, District of Columbia, United States of America; University of West Virginia, School of Medicine, Morgantown, West Virginia, United States of America; University of Rhode Island, College of Pharmacy, Kingston, Rhode Island

## Abstract

**Introduction:** Medication Regimen Complexity (MRC) refers to the combination of medication classes, dosages, and frequencies. The relationship between MRC and clinical outcomes in the intensive care unit (ICU) has not been examined. The objective of this study is to examine the association between MRC scoring tools and their utility in predicting clinical outcomes.

**Methods:** We conducted a retrospective cohort study that includes 322 adult patients admitted and stayed (>24 hours) to the ICU between February 1, 2020, and August 30, 2020 in a community-based, teaching hospital in Rhode Island. Medication complexity was assessed using two MRC scoring tools: MRC Index (MRCI) and the MRC in ICU (MRCICU). We used a multivariable logistic regression model to identify the association between MRC scores and clinical outcomes and to predict the clinical outcomes.

**RESULTS:** Among the 317 patients included in the study (55.2% men with a median age of 62 [IQR: 51-75] years). Higher MRC scores (i.e., > 63 MRCI or > 6 MRCICU) were associated with increased mortality (14% and 15%), longer ICU length of stay (LOS)(30% and 34%), and need for MV (24% and 28%). MRCICU scores at 24 hours were found to be a significant risk factor in all clinical outcomes (ICU mortality, LOS, and MV) with Odds Ratio (ORs) of 1.12 (95% CI: 1.06-1.19), 1.17 (1.1-1.24), and 1.21 (1.14-1.29), respectively. In the prediction models, both MRCI and MRCICU models performed similarly (AUC: 0.88 [0.75-0.97] and 0.88 [0.76-0.97] in predicting mortality. The Medication Model included 15 medication classes outperformed others (AUC: 0.82 [0.71-0.93] in predicting ICU LOS and the MRCICU model outperformed others (AUC: 0.87 [0.77-0.96]) in predicting the need for MV.

**CONCLUSION:** MRC scores are associated with poorer clinical outcomes and improves the prediction of poorer clinical outcomes which will support clinicians to prescribe safer therapies.

## Introduction

Medication regimen complexity (MRC) refers to multiple features of a patient’s medication drug regimen rather than absolute number of medications consumed per day[1]. MRC incorporates features such as the number of agents, dosages, administration time intervals, and additional instructions (i.e., take on an empty stomach)[2–4]. An increase in MRC burden has been associated with poorer medication noncompliance and caregiver quality of life measures, as well as an increase in healthcare resource utilization[5,6]. Critically ill patients are at significant risk of higher MRC due to the severity of illness, management of multiple chronic conditions, and the complex pharmacotherapies prescribed. It has been estimated that the average critically ill adult may receive up to 13 medications per day and the incidence of experiencing an adverse drug event has been estimated to greater than 25% lending to substantial morbidity and mortality[7–11]. Therefore, examining only the quantity of medications administered may not accurately describe the complex and intricate nature of the critical care medication regimens.

Numerous methods have been used to quantify the complexity of medication regimens. Yet, the most commonly utilized and validated objective scoring tool is the 65-item, weighted MRC index (MRCI) which has been developed for outpatient use[12–16]. MRCI has been used to evaluate conditions such as neurological impairment in children, hypertension, diabetes, and chronic kidney disease in adults[17– 21]. In the intensive care unit (ICU) population, the MRC in ICU (MRCICU) scoring tool was developed and revised (modified MRCICU) in 2019[22,23]. The modified MRCICU is the first validated quantitative weighted scoring tool intended to predict clinical outcomes (i.e., ICU mortality, length of stay (LOS), and need for mechanical ventilation (MV))[24,25]. In a previous study, we have developed a novel proof-of-concept method demonstrating improvement in the prediction of patient outcomes by incorporating the MRCICU score into the previously established Acute Physiology and Chronic Health Evaluation (APACHE II) scoring tool[26]. To date, no study has assessed the association between these two MRC scoring tools and clinical outcomes within the critical care setting.

In this study, we developed two custom MRC scoring algorithms and several statistical and prediction models using the MRCI and MRCICU tools to gain insight into how MRC impacts clinical outcomes (i.e., ICU mortality, LOS, and need for MV). We aimed to (1) examine how MRC scores correlate with clinical characteristics (2) test a hypothesis that MRC scores influence clinical outcomes (3) determine the utility of MRC scores in predicting clinical outcomes for use in practice.

## Methods

### Study Design

This was a single center, retrospective cohort study of 322 patients enrolled into the ICU in a 220-bed community hospital in Providence, Rhode Island, USA between February 1, 2020, and August 30, 2020. Due to the retrospective nature of the data, informed consent was not deemed necessary as all patient data was de-identified prior to use. The study was granted exemption by the Human Research Review Committee Roger Williams Medical Center Institutional Review Board (IRB: 00000058) and University of Rhode Island Institutional Review Board (IRB: 00000559). All adult patients admitted to the medical-surgical intensive care were included if the following criteria were met: ≥ 18 years of age and admission to the ICU > 24 hours. Demographic, vital signs, laboratories, medication classes, and MV data were collected for each patient from the electronic health records (EHR) (Table S1). Five patients were excluded due to extensive LOS (> 40 days) and omitted laboratory values. A total of 317 patients were included in the final analysis.

### MRC scoring tools

In this paper, mortality and LOS refer to ICU setting. MRC can be defined by the number of drugs and their dosing frequency. Two medication scoring indexes, MRCI and the MRCICU, have been developed for outpatient and acute care use, respectively. The total MRCI score is the weighted sum of 3 sections (dosage form, dosing frequency, and additional administration information), in which a higher score reflects higher MRC. In computing the MRCI, both scheduled and “as needed” medications and supplements were considered^1^.

Conversely, the MRCICU is a 39-item weighted critically ill scoring tool comprised of specific medications and classes (i.e., vancomycin-3 points, continuous intravenous saline-1 point)[27]. The MRCICU scoring table and individually assigned medication weights are presented elsewhere. The score has undergone validation testing for both internal and external validity[27]. Recently, we developed a proof-of-concept model using machine learning (ML) methods to validate the MRCICU score for improving the prediction of inpatient mortality[26]. Although a MRCICU score can be determined at any time during ICU admission, historical evaluation daily at 24-hour intervals have been most commonly utilized and have been applied to the present study. These two scores were custom coded for each patient during each 24-hour interval.

### Definitions

We defined cutoff values for both MRC scores based upon their distribution after 24 hours of ICU admission. A ‘high’ MRCI cohort was defined as a score of ≥ 63 and a ‘high’ MRCICU cohort of ≥ 6. We utilized binary variables for LOS = 0 when LOS was < 48 hours and LOS = 1 when LOS > 48 hours. The need for MV was defined using a binary variable of (MV = 0, not mechanically ventilated and MV = 1, mechanically ventilated).

### Statistical analysis

Descriptive statistics were used to describe the study population where continuous values were represented using means and interquartile ranges (IQR). Categorical variables were described using frequencies and proportions. Comparisons between survivor and non-survivor and low or high MRC scoring cohorts were performed using Student’s *t*, Chi-square (χ^2^), or Fisher’s exact tests. Physiological and clinical characteristics were compared among the survivor and non-survivor cohorts. To account for severity of illness, we included the following scoring tools: Simplified Acute Physiology Score (SAPS II), APACHE II, and Charlson Comorbidity Index (CCI). MRCI and MRCICU were analyzed for mortality, need for MV, and SARS-CoV-2 (COVID-19) infection using the Wilcoxon signed rank test. Four multivariable logistic regression models were utilized to examine the association between risk factors and clinical outcomes (i.e., mortality, LOS, and need for MV). The four models were: Model I: Demographics, APACHE II, SAPS II, CCI, and 15 drug classes, Model II: Demographics, MRCI_24hours, MRCI_48hours, CCI, and drug classes, Model III: Demographics, MRCICU_24hours, MRCICU_48hours, CCI, and drug classes, and Model IV: all variables. In the LOS models, we excluded MRCI and MRCICU values at 48 hours, as our threshold for binary values was 48 hours of ICU admission. Significant predictors (*p* < 0.05) were selected for each model using a stepwise forward selection method. The best fit models were selected using Akaike information criterion (AIC). To further assess variable selection, we used a L1 penalization technique (LASSO). LASSO allows for a more restrictive parameter selection that is minimally influenced by multicollinearity. Demographic variables included were age, sex, height, weight, body mass index (BMI), and race. Odds Ratios (OR) were calculated for each outcome of interest. All analyses were conducted using R, Version 4.0.0 (R Project for Statistical Computing), *glm, glmnet, and ggplot2 R* packages were used[28–30].

### Prediction model development

To test the predictability of MRC scores for mortality, LOS, and need for MV, seven logistic regression prediction models were constructed without any variable selection. A ‘no imputation’ approach was used when preparing the data for the prediction model. We assessed correlated variables using the Pearson correlation coefficient. The SAPS II severity score was used in the prediction models due to a high correlation with the APACHE II classification system (S1 Fig). The prediction accuracy was evaluated using the area under the receiver operating characteristic (AUC) curve. An AUC of at least 0.7 was regarded as acceptable. We applied a ‘leave-one-out’ cross-validation method with 10,000 repetitions and AUC was selected as an overall performance measure. Additionally, sensitivity and specificity analyses were included for the three outcomes. Furthermore, all prediction models recorded variable importance rankings for each outcome.

## Results

### Demographic and clinical characteristics

Of the 317 patients included in the analysis, most were male (175 patients [55.2%]); and White (205 patients [65%]), with a median (interquartile range [IQR]) age of 62 (51-75) years. A total of 77% patients survived, 51% had a LOS > 48 hours, and 31% required MV. Vital signs, serum electrolytes, and blood cell values were similar among the survivors and non-survivor’s cohorts. Conversely, serum BUN, creatinine values, and eGFR’s were significantly worse in the non-survivor cohort (25.7mg/dL, 1.5mg/dL, 71.2 mL/min/1.73m^2^ vs 38.7mg/dL, 1.8mg/dL, 50.3 ml/min/1.73m^2^), respectively. Non-survivors had a significantly longer duration on MV (147.2 hours vs. 34.6 hours) and prolonged LOS (191.4 hours vs. 87.4 hours) when compared to the survivor cohort. There was a high prevalence among both cohorts with acute respiratory failure with hypoxia (125 [39.4%], COVID infection (52 [16.4%]), lactic acidosis (101 [31.9%], kidney failure (96 [30.3%]), and acute myocardial infarction (78 [24.6%]) (**Table 1**).

**Table 1.**
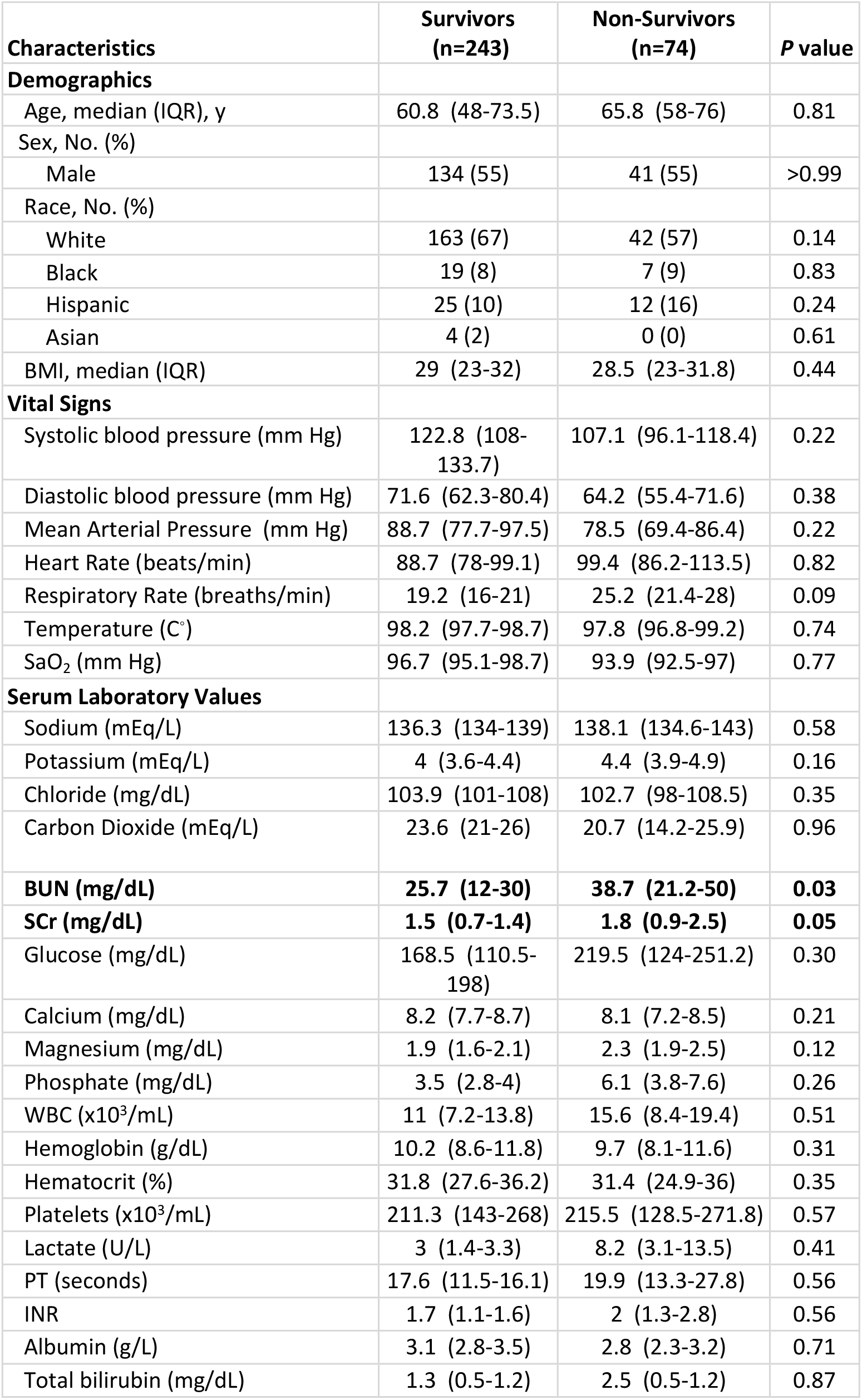
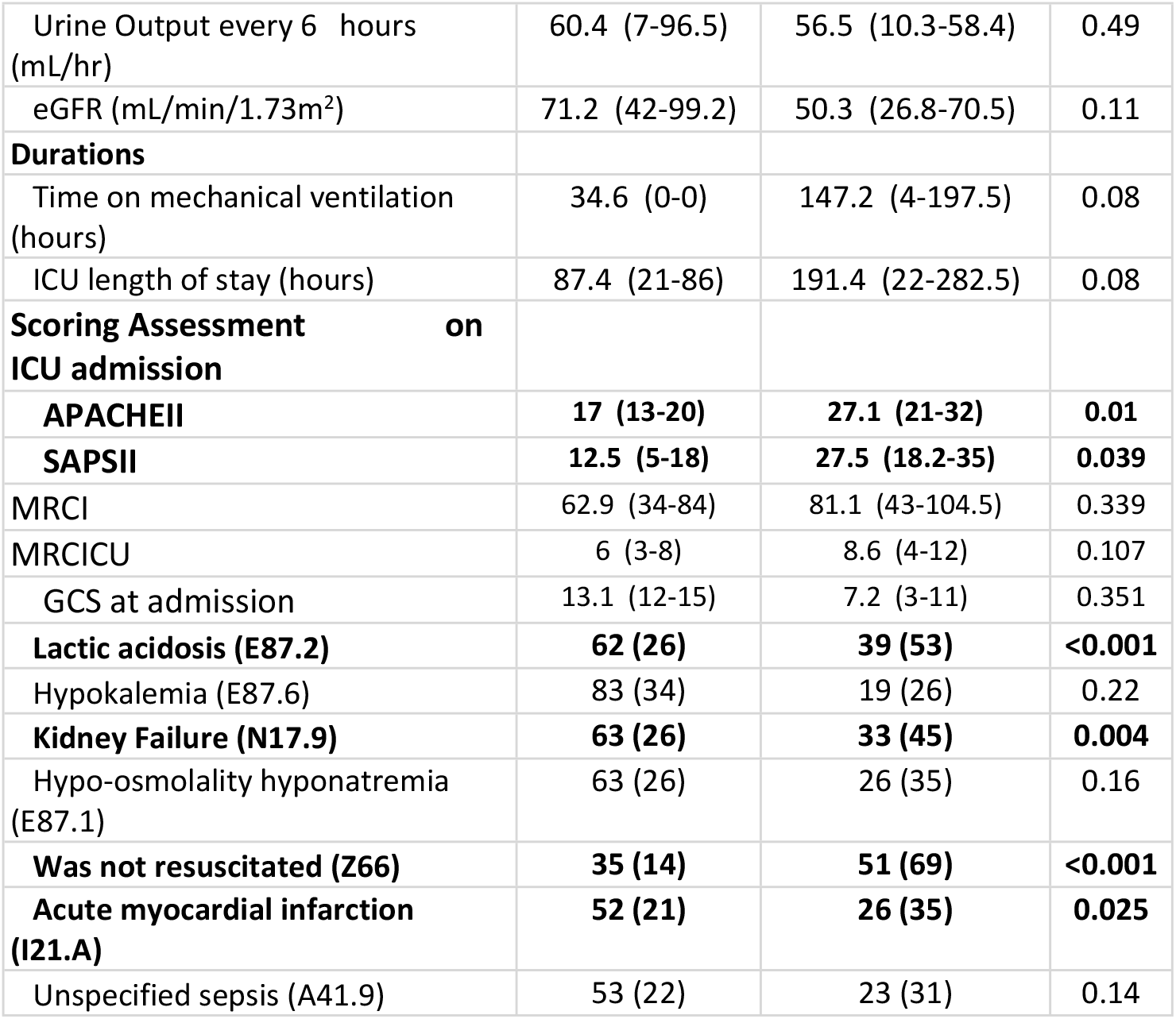
Comparison of survivors and non-survivors cohorts. Comparing the demographics and clinical characteristics between survivors and non-survivors

| Characteristics | Survivors<br>(n=243) | Non-Survivors<br>(n=74) | P value |
| --- | --- | --- | --- |
| **Demographics** |  |  |  |
| Age, median (IQR), y | 60.8 (48-73.5) | 65.8 (58-76) | 0.81 |
| Sex, No. (%) |  |  |  |
| Male | 134 (55) | 41 (55) | >0.99 |
| Race, No. (%) |  |  |  |
| White | 163 (67) | 42 (57) | 0.14 |
| Black | 19 (8) | 7 (9) | 0.83 |
| Hispanic | 25 (10) | 12 (16) | 0.24 |
| Asian | 4 (2) | 0 (0) | 0.61 |
| BMI, median (IQR) | 29 (23-32) | 28.5 (23-31.8) | 0.44 |
| **Vital Signs** |  |  |  |
| Systolic blood pressure (mm Hg) | 122.8 (108-133.7) | 107.1 (96.1-118.4) | 0.22 |
| Diastolic blood pressure (mm Hg) | 71.6 (62.3-80.4) | 64.2 (55.4-71.6) | 0.38 |
| Mean Arterial Pressure (mm Hg) | 88.7 (77.7-97.5) | 78.5 (69.4-86.4) | 0.22 |
| Heart Rate (beats/min) | 88.7 (78-99.1) | 99.4 (86.2-113.5) | 0.82 |
| Respiratory Rate (breaths/min) | 19.2 (16-21) | 25.2 (21.4-28) | 0.09 |
| Temperature (C°) | 98.2 (97.7-98.7) | 97.8 (96.8-99.2) | 0.74 |
| SaO<sub>2</sub> (mm Hg) | 96.7 (95.1-98.7) | 93.9 (92.5-97) | 0.77 |
| **Serum Laboratory Values** |  |  |  |
| Sodium (mEq/L) | 136.3 (134-139) | 138.1 (134.6-143) | 0.58 |
| Potassium (mEq/L) | 4 (3.6-4.4) | 4.4 (3.9-4.9) | 0.16 |
| Chloride (mg/dL) | 103.9 (101-108) | 102.7 (98-108.5) | 0.35 |
| Carbon Dioxide (mEq/L) | 23.6 (21-26) | 20.7 (14.2-25.9) | 0.96 |
| **BUN (mg/dL)** | **25.7 (12-30)** | **38.7 (21.2-50)** | **0.03** |
| **SCr (mg/dL)** | **1.5 (0.7-1.4)** | **1.8 (0.9-2.5)** | **0.05** |
| Glucose (mg/dL) | 168.5 (110.5-198) | 219.5 (124-251.2) | 0.30 |
| Calcium (mg/dL) | 8.2 (7.7-8.7) | 8.1 (7.2-8.5) | 0.21 |
| Magnesium (mg/dL) | 1.9 (1.6-2.1) | 2.3 (1.9-2.5) | 0.12 |
| Phosphate (mg/dL) | 3.5 (2.8-4) | 6.1 (3.8-7.6) | 0.26 |
| WBC (x10<sup>3</sup>/mL) | 11 (7.2-13.8) | 15.6 (8.4-19.4) | 0.51 |
| Hemoglobin (g/dL) | 10.2 (8.6-11.8) | 9.7 (8.1-11.6) | 0.31 |
| Hematocrit (%) | 31.8 (27.6-36.2) | 31.4 (24.9-36) | 0.35 |
| Platelets (x10<sup>3</sup>/mL) | 211.3 (143-268) | 215.5 (128.5-271.8) | 0.57 |
| Lactate (U/L) | 3 (1.4-3.3) | 8.2 (3.1-13.5) | 0.41 |
| PT (seconds) | 17.6 (11.5-16.1) | 19.9 (13.3-27.8) | 0.56 |
| INR | 1.7 (1.1-1.6) | 2 (1.3-2.8) | 0.56 |
| Albumin (g/L) | 3.1 (2.8-3.5) | 2.8 (2.3-3.2) | 0.71 |
| Total bilirubin (mg/dL) | 1.3 (0.5-1.2) | 2.5 (0.5-1.2) | 0.87 |
| Urine Output every 6 hours (mL/hr) | 60.4 (7-96.5) | 56.5 (10.3-58.4) | 0.49 |
| eGFR (mL/min/1.73m<sup>2</sup>) | 71.2 (42-99.2) | 50.3 (26.8-70.5) | 0.11 |
| **Durations** |  |  |  |
| Time on mechanical ventilation (hours) | 34.6 (0-0) | 147.2 (4-197.5) | 0.08 |
| ICU length of stay (hours) | 87.4 (21-86) | 191.4 (22-282.5) | 0.08 |
| **Scoring Assessment on ICU admission** |  |  |  |
| **APACHEII** | **17 (13-20)** | **27.1 (21-32)** | **0.01** |
| **SAPSII** | **12.5 (5-18)** | **27.5 (18.2-35)** | **0.039** |
| MRCI | 62.9 (34-84) | 81.1 (43-104.5) | 0.339 |
| MRCICU | 6 (3-8) | 8.6 (4-12) | 0.107 |
| GCS at admission | 13.1 (12-15) | 7.2 (3-11) | 0.351 |
| **Lactic acidosis (E87.2)** | **62 (26)** | **39 (53)** | **<0.001** |
| Hypokalemia (E87.6) | 83 (34) | 19 (26) | 0.22 |
| **Kidney Failure (N17.9)** | **63 (26)** | **33 (45)** | **0.004** |
| Hypo-osmolality hyponatremia (E87.1) | 63 (26) | 26 (35) | 0.16 |
| **Was not resuscitated (Z66)** | **35 (14)** | **51 (69)** | **<0.001** |
| **Acute myocardial infarction (I21.A)** | **52 (21)** | **26 (35)** | **0.025** |
| Unspecified sepsis (A41.9) | 53 (22) | 23 (31) | 0.14 |

### Clinical characteristics between MRC cohorts

In the MRCI cohort, lower vital sign values (SBP, DBP, and MAP) were found to be significant within the higher MRC scoring group. Serum laboratory indices including phosphate, lactate, and albumin varied significantly among the MRCI cohort. Higher MRCI scores correlated with increased patient acuity when compared to lower MRCI scoring groups. The comorbidities hypo-osmolality, acute myocardial infarction (AMI), and unspecified sepsis were significant among the MRCI and MRCICU cohorts. **(S2 and S3 Tables)**.

When compared to the non-survivor cohort, the survivor cohort had significantly lower MRCI, MRCICU, APACHE II, and SAPS II scores. In the COVID-19 infected cohort, APACHE II and SAPS II had significantly lower median values when compared to the non-COVID infected group. Further, MRCI and MRCICU scores were significantly higher in the mechanically ventilated cohort **(Fig 1)**. When analyzing age distribution by decade of life among different comorbidity severity indexes (i.e., Charlson, APACHE II, SAPS II) patients greater than sixty years of age were associated with the highest severity index scores. **(S2 Fig)**. The top 5 medication classes prescribed among the high scoring MRC cohorts (MRCI and MRCICU) were intravenous fluids (normal saline, 36% and 35%), gastrointestinal agents (pantoprazole, 29% and 32%), analgesics (acetaminophen, 26% and 26%), electrolytes (potassium chloride, 24% and 26%) and anti-infectives (piperacillin/tazobactam, 23% and vancomycin, 27%). When incorporating the severity of illness scoring tools (APACHE II and SAPS II) with MRCI and MRCICU scores patients with higher MRC scores (i.e., > 63 MRCI and > 6 MRCICU) were associated with increased mortality (14% and 15%), longer LOS (i.e., > 48 hours; 30% and 34%), and an increased need for MV (24% and 28%), respectively **(S3 Fig)**.

**Fig 1.**
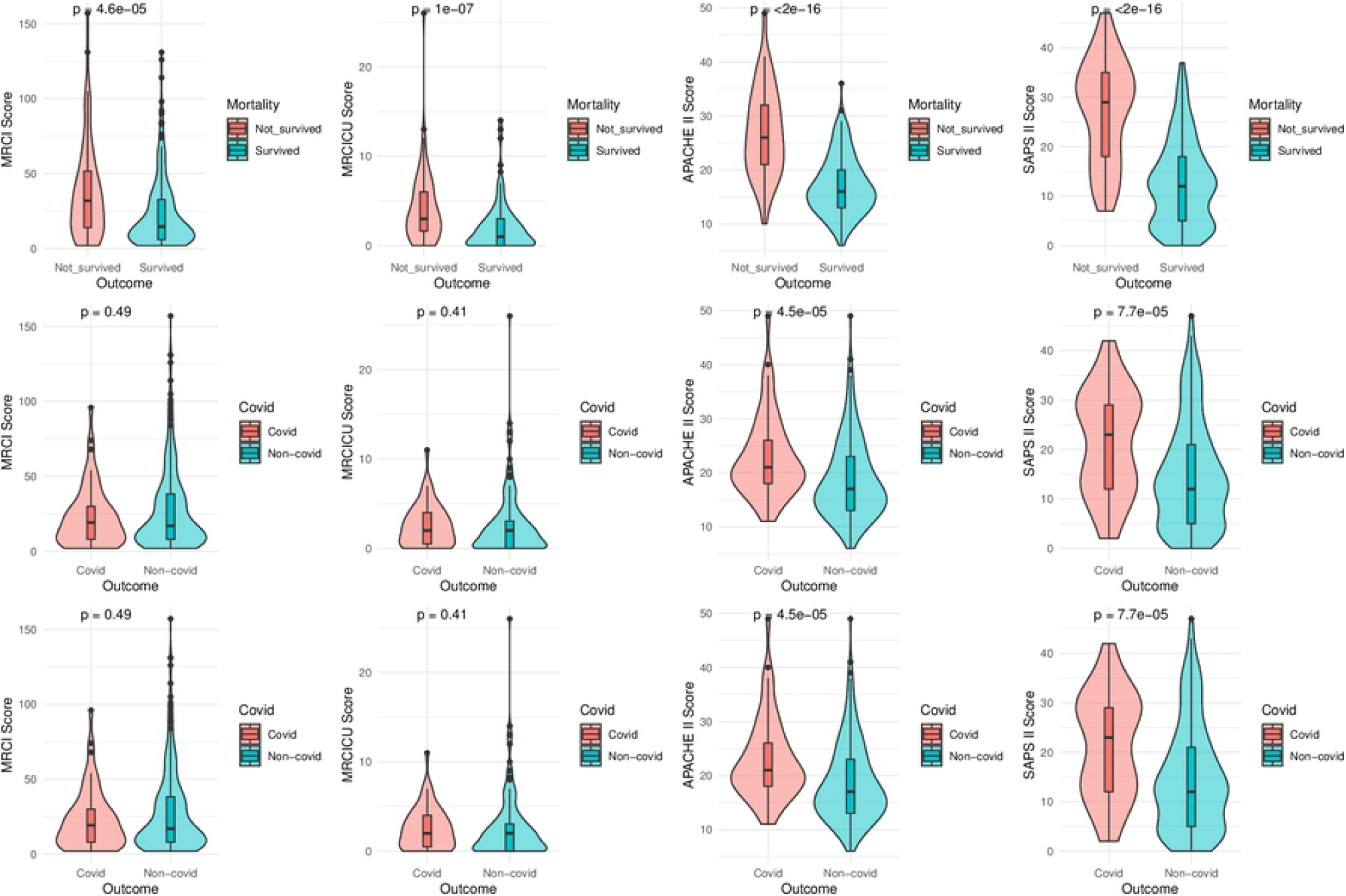
Comparisons among the severity score and MRC scores. Comparison of severity of illness scores Acute Physiology and Chronic Health Evaluation (APACHE II), Simplified Acute Physiology Score (SAPS) II, Medication Regimen Complexity Index (MRCI), and Medication Regimen Complexity in Intensive Care Unit (MRCICU) between survivor and non-survivor (top

### Associations between MRC scores and clinical outcomes

At admission, SAPS II was significant for all three outcomes: mortality (OR: 1.12 (1.07-1.18), LOS (OR: 1.04 (1.0-1.11), need for MV (OR: 1.17 (1.13-1.21) (**Table 2**), respectively. When incorporating only MRCI scores into the model, (Model II), MRCI scores at 24 hours was a significant predictor in all outcomes with ORs of 1.01 (95% CI: 1.0-1.02), 1.01 (1.0-1.02), and 1.01 (1.01-1.02) for mortality, LOS, and need of MV, respectively. Further, MRCI scores at 48 hours were found to be significant risk factors for mortality and need for MV (**Table 2**). MRCICU scores at 24 hours were significant risk factors in all outcomes in Model III with ORs of 1.12 (95% CI: 1.06-1.19), 1.17 (1.1-1.24), and 1.21 (1.14-1.29) for mortality, LOS, and need of MV, respectively. Complimentary results from the LASSO model confirmed the risk factor selection trends (**S4 Table**). The use of vasopressors was found to be a significant risk factor for all outcomes in the models. When evaluating morality, the use of paralytic agents was significant with an (OR: 3.38 (1.09-11.11). The use of anti-infectives, anticoagulants, and cardiovascular agents were significantly associated with prolonged LOS. Lastly, the use of analgesics, sedatives, psychiatric, cardiovascular, and pulmonary agents were significant risk factors for the need of MV.

**Table 2.** Identifying the risk factors for ICU mortality, length of stay (LOS) and need for mechanical ventilation (MV). List of selected variables using stepwise selection method for the four logistic regression models and their associations with mortality, LOS, and need for MV.

| Selected features | Mortality<br>OR (95% CI) | p-value | Length of ICU<br>Stay<br>OR (95% CI) | p-value | Mechanical<br>Ventilation<br>OR (95% CI) | p-value |
| --- | --- | --- | --- | --- | --- | --- |
| **Model I** |  |  |  |  |  |  |
| Age | 1.02 (1.0 - 1.05) | 0.12 | 1.02 (1.0-1.03) | 0.05 | - | - |
| BMI | - | - | 1.02 (1.0-1.05) | 0.11 | - | - |
| White | - | - | 0.59 (0.35-0.98) | - | - | - |
| Hispanic | 6.11 (2.84 - 13.85) | <0.001 | - | - | - | - |
| **SAPS II at admission** | **1.12 (1.07-1.18)** | **0.001** | **1.04 (1- 1.11)** | **0** | **1.17 (1.13-1.21)** | **0** |
| **APACHE II at admission** | **1.13 (1.05- 1.21)** | **0.002** | 0.92 (0.88- 0.98) | 0.15 | - | - |
| CCI | - | - | 1.13 (1.0-1.32) | 0.14 | - | - |
| **Model II** |  |  |  |  |  |  |
| Age | 1.03 (1.02- 1.05) | <0.001 | 1.01 (1.0- 1.03) | 0.09 | - | - |
| Height | - | - | 1.14 (1.01- 1.35) | 0.11 | 1.02 (1.04- 1.46) | 0.03 |
| Weight | - | - | 0.95 (0.89- 1.01) | 0.09 | 0.95 (0.89-1.01) | 0.1 |
| BMI | - | - | 1.17 (1.0- 1.41) | 0.06 | 1.18 (1.0-1.42) | 0.07 |
| White | - | - | 0.56 (0.33-0.93) | 0.03 | - | - |
| Hispanic | 5.74 (3.15- 10.79) | 0 | - | - | 1.84 (1.11-3.07) | 0.02 |
| **MRCI score at 24 hours** | **1.01 (1- 1.02)** | **0.003** | **1.01 (1.0-1.02)** | **0** | **1.01 (1.01-1.02)** | **0** |
| **MRCI score at 48 hours** | **1.01 (1- 1.02)** | **0.004** | - |  | **1.01 (1.0-1.02)** | **0.03** |
| CCI | - | - | 1.13 (1.0-1.32) | 0.13 | - | - |
| **Model III** |  |  |  |  |  |  |
| Age | 1.03 (1.02- 1.05) | <0.001 | 1.01 (1.0- 1.03) | 0.1 | - | - |
| White | - | - | 0.59 (0.35-0.99) | 0.04 | - | - |
| Hispanic | 6.44 (3.45- 12.41) | 0 | - | - | 2.21 (1.29- 3.85) | 0.004 |
| **MRC ICU at 24 Hours** | **1.12 (1.06- 1.19)** | **0** | **1.17 (1.1- 1.24)** | **<0.001** | **1.21 (1.14- 1.29)** | **<0.001** |
| **MRC ICU at 48 hours** | **1.1 (1.04-1.17)** | **0.002** | 1.05 (1.0-1.11) | 0.7 | **1.11 (1.05- 1.18)** | **0.001** |
| CCI | - | - | 1.14 (1.0-1.34) | 0.1 | 0.84 (0.65- 1.07) | 0.155 |
| **Model IV** |  |  |  |  |  |  |
| Age | 1.04 (1.01- 1.08) | 0.017 | - | - | - | - |
| Hispanic | 6.23 (2.55- 16.41) | 0 | - | - | - | - |
| **SAPS II at admission** | **1.09 (1.04- 1.15)** | **0.001** | **1.04 (1- 1.08)** | **0.072** | **1.17 (1.11- 1.23)** | **0** |
| **APACHE II at admission** | **1.14 (1.05- 1.23)** | **0.002** | **0.94 (0.89- 1)** | **0.038** | - | - |
| **MRCI at 24 hours** | - | - | **0.99 (0.98- 1)** | **0.134** | **0.97 (0.95- 0.99)** | **0.001** |
| **MRCICU at 24 hours** | - | - | **1.1 (1- 1.22)** | **0.05** | **1.3 (1.13- 1.51)** | **0** |
| CCI | 0.79 (0.6- 1.03) | 0.093 | - | - | 0.77 (0.58- 1) | 0.052 |
| Anti-infectives | - | - | 2.27 (1.15- 4.57) | 0.019 | - | - |
| Anticoagulants | 0.38 (0.1- 1.42) | 0.139 | 2.26 (1.02- 5.29) | 0.05 | - | - |
| Psychiatric agents | - | - | 1.8 (0.96- 3.36) | 0.065 | 2.52 (1.01- 6.64) | 0.05 |
| Pulmonary agents | - | - | - | - | 3.14 (1.34- 7.66) | 0.01 |
| Cardiovascular agents | - | - | 2.81 (1.46- 5.53) | 0.002 | 0.4 (0.15- 0.98) | 0.05 |
| Diuretics | - | - | 3.35 (1.74- 6.62) | 0 | - | - |
| Analgesics sedatives | - | - | - | - | 6.96 (1.73- 36.07) | 0.012 |
| Vasopressors | 5.55 (2.12- 15.26) | 0.001 | 3.49 (1.63- 7.75) | 0.002 | 5.75 (2.4- 14.48) | 0 |
| Paralytic agents | 3.38 (1.09- 11.11) | 0.039 | - | - | - | - |
| Vitamins | - | - | 1.63 (0.88- 3.03) | 0.122 | 0.25 (0.09- 0.62) | 0.004 |
| Others | 2.54 (1.08- 6.15) | 0.034 | - | - | - | - |

The final models of logistic regression are reported using odds ratio (OR) and 95% confidence interval of risk factors for logistic regression. If the variable was not selected, the cell marked with "-". Bold ORs for logistic regression are significant. Model I: Demographics, Acute Physiology and Chronic Health Evaluation (APACHE II), Simplified Acute Physiology Score (SAPS II), Charlson comorbidity index (CCI), and drug classes, Model II: Demographics, Medication Regimen Complexity Index (MRCI\_24hours), MRCI\_48hours, CCI, and drug classes, Model III: Demographics, Medication Regimen Complexity in Intensive Care Unit (MRCICU\_24hours), MRCICU\_48 hours, CCI, and drug classes, and Model IV: all variables.

### Role of MRC scores in prediction of clinical outcomes

The Admission Model was found to be the best model (AUC: 0.88 [95% CI: 0.77-0.97]) to predict mortality (**Table 3**). However, models MRCI & SAPS II (AUC: 0.88 [0.75-0.97]) and MRCICU & SAPS (AUC: 0.88 [0.76-0.97]) performed similarly (**Fig 2**). In the MRCI & SAPS II Model, MRCI scores at 24 and 48 hours were identified as the top variables of importance when predicting mortality (**Fig 3**). Further, vasopressors were the most important variable to predict mortality within the Medication Model.

**Table 3.**
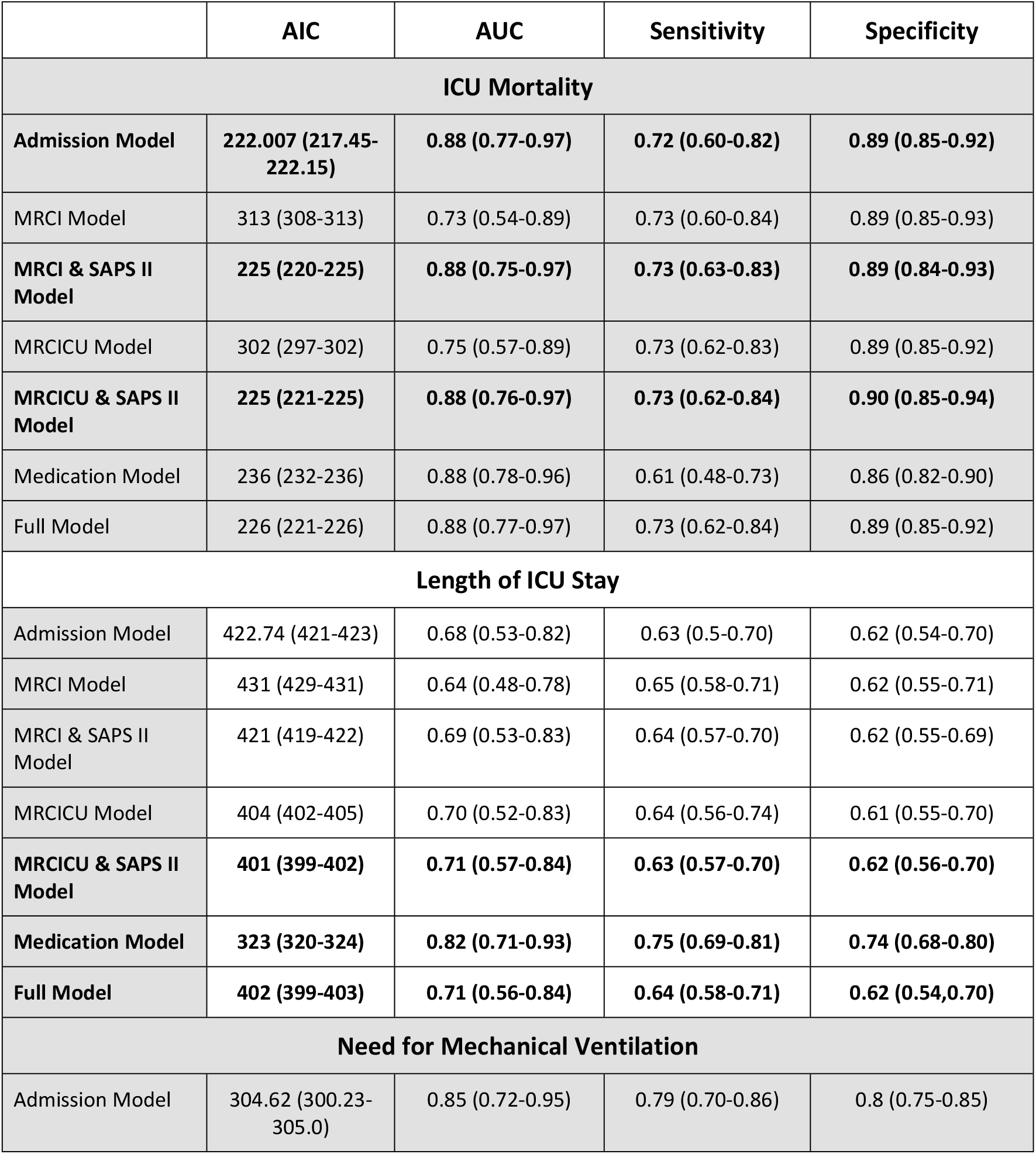
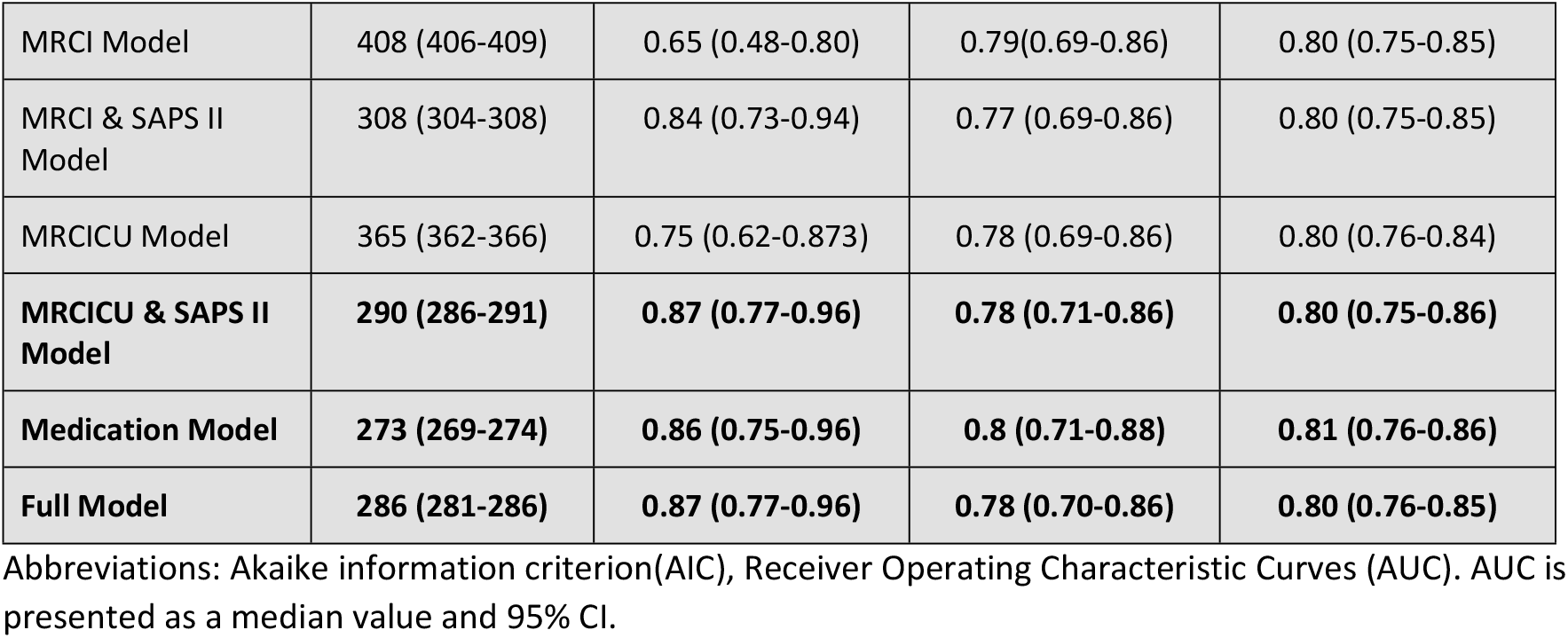
Comparison of the prediction models. Prediction evaluation for ICU mortality, LOS, and need for mechanical ventilation. (The best 3 prediction results are in noted in bold font and demographic variables are included in each of the models).

**Fig 2.**
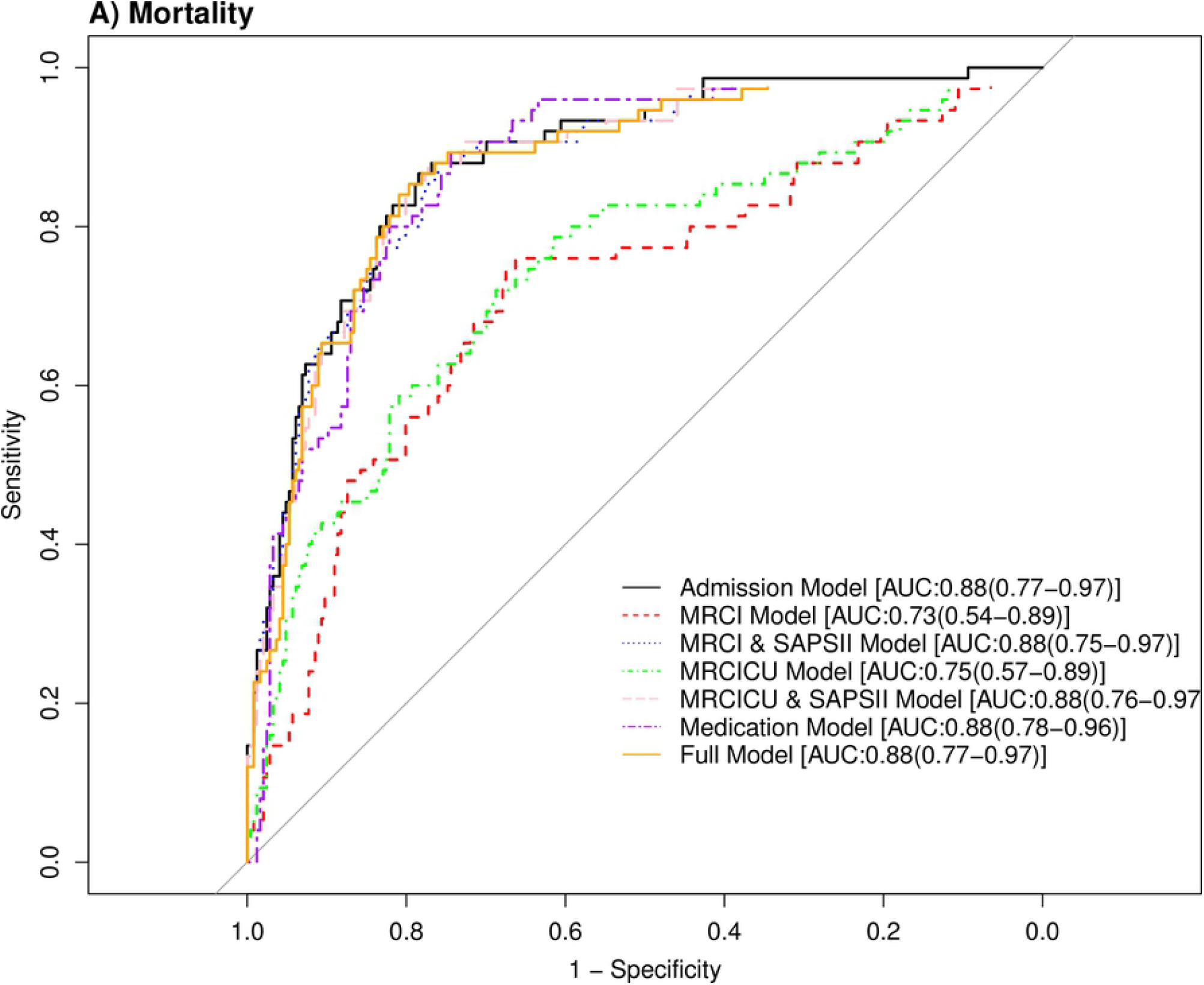
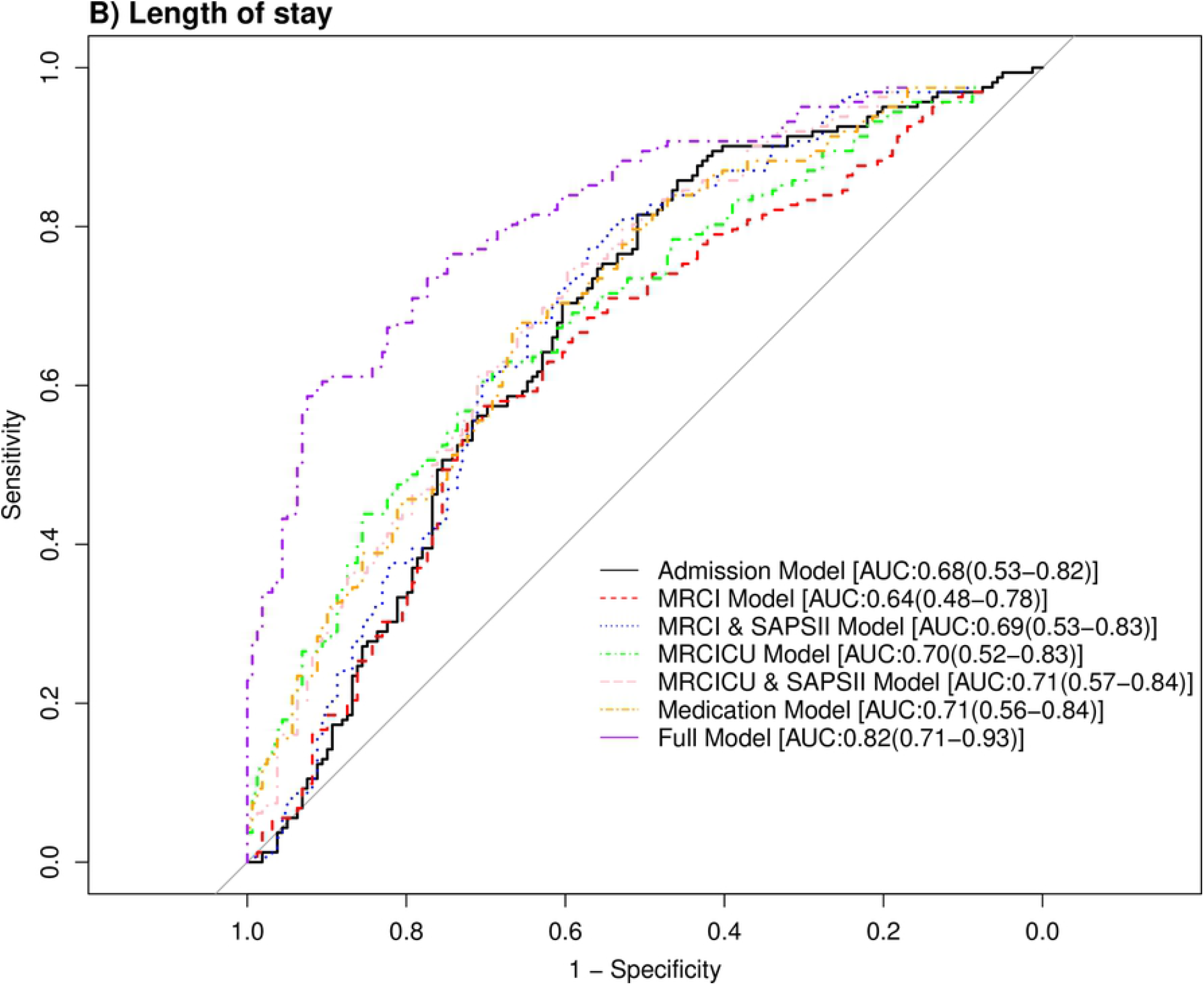
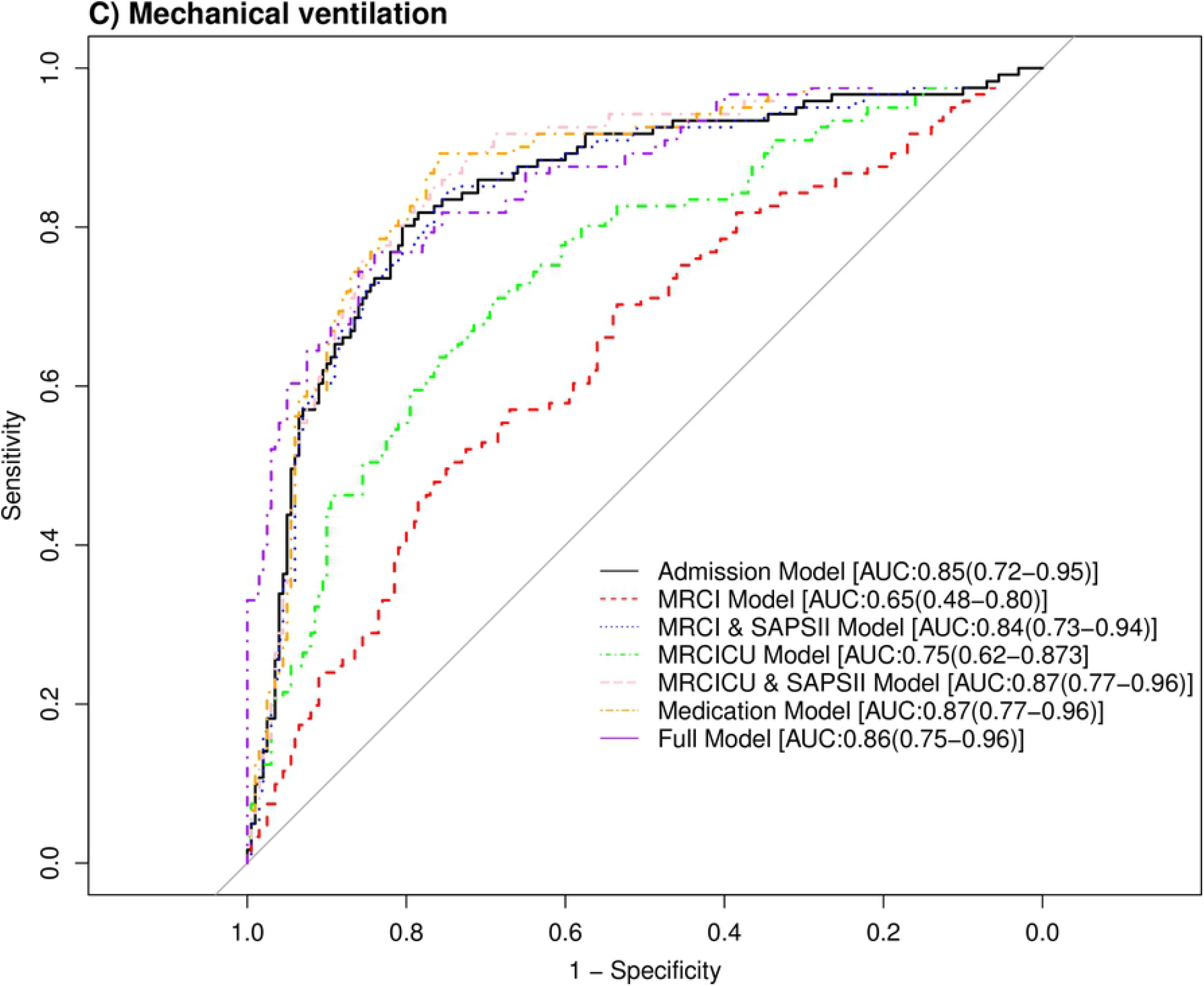
Performance of the prediction models for predicting the clinical outcomes. Receiver Operating Characteristic Curves (AUC) for (A) ICU mortality, (B) ICU length of stay, and (C) need for mechanical ventilation.

**Fig 3.**
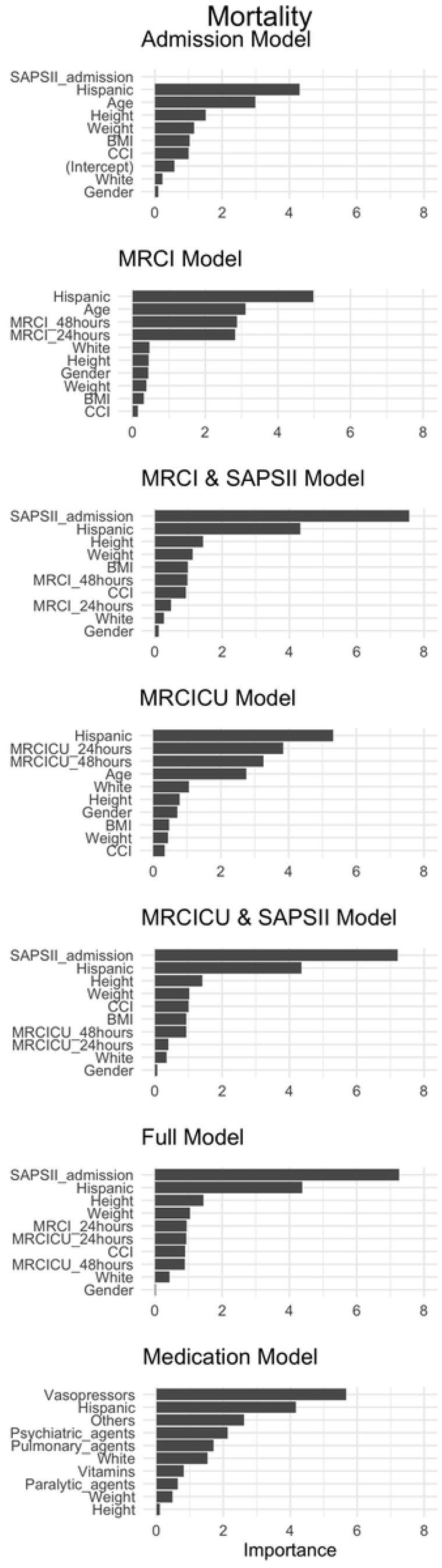
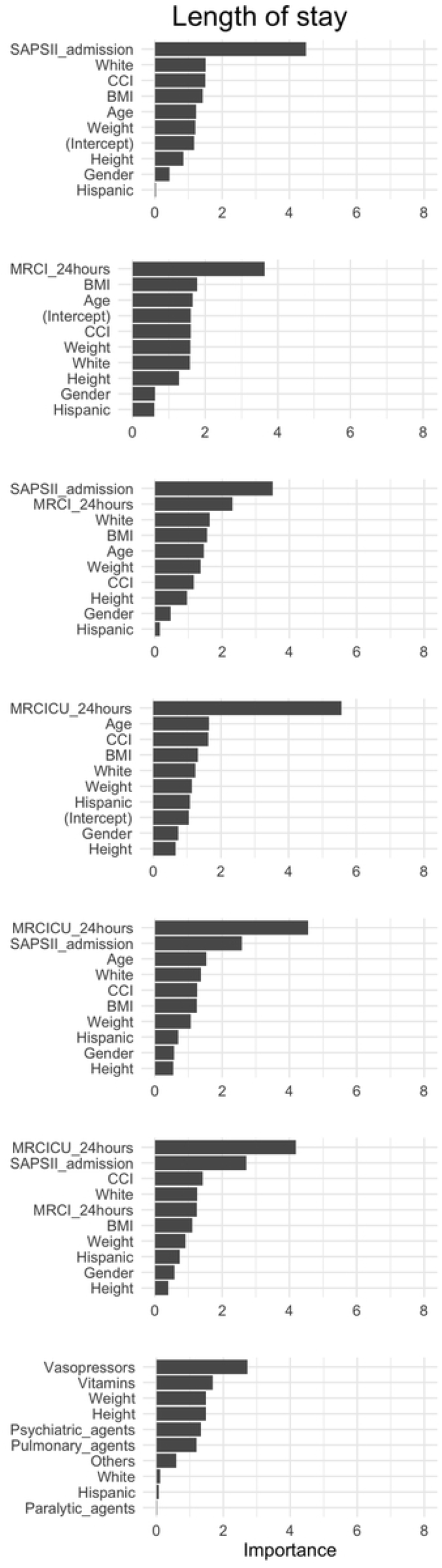
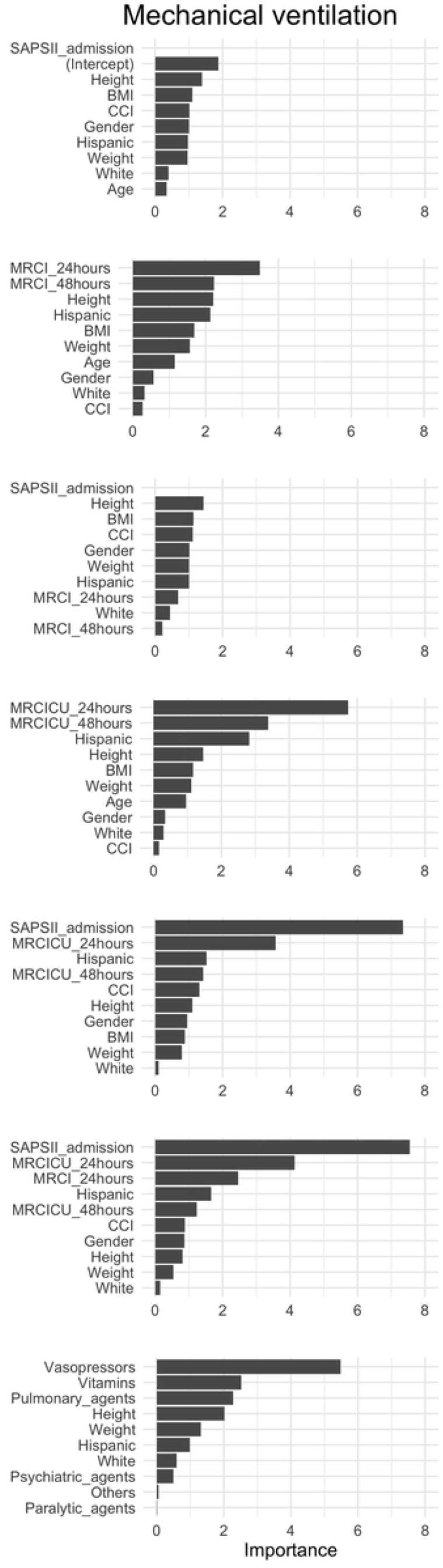
Variable importance of the prediction models. Top 10 variables of importance within the 7 prediction models: ICU mortality (*left* panel), length of ICU stay (*middle* panel), and need for mechanical ventilation (*right* panel).

When predicting LOS, the Medication Model (AUC: 0.82 [0.71-0.93]) outperformed all other models. Vasopressors and psychiatric agents were among the top 5 important variables to predict LOS. Further, MRC scores at 24 hours and 48 hours were selected in the top 10 variable importance list for models including MRC scores (i.e., MRCI & SAPS II and, MRCICU & SAPS II).

When predicting the need for MV, MRCICU & SAPS II (AUC: 0.87 [0.77-0.96]) outperformed all other models. SAPS II and MRCICU at 24 and 48 hours were among the top important variables to predict the need for MV. Lastly, vasopressors and pulmonary agents were among the top 5 medication classes identified when predicting the need for MV.

## Discussion

### Key findings

In this study, we found that MRC scores varied greatly and regression analysis confirmed that MRC scores are strongly associated with all outcomes: mortality, LOS, and need for MV. Secondly, higher MRC scores were associated with hemodynamic instability and higher APACHE II scores. Survivors had significantly lower MRCI, MRCICU, APACHE II, and SAPS II scores. Thirdly, incorporating MRC scores improved the prediction in all outcomes. The utility of MRC scores and impact on clinical outcomes in the critical care setting remains largely unknown. To the best of our knowledge, this is the first study to examine these relationships. Previous studies suggest inconsistent findings when investigating MRC scores with medication nonadherence and hospitalization in outpatient settings[31–35]. In the ICU, MRC has been correlated with patient acuity and mortality ^18^. Consistent with previously published literature, our study revealed non-survivors had poorer renal function, increased time on MV, and extended ICU LOS[36–39]. Notably, when predicting LOS, MRC scores at 24 hours were considered to be a significant prediction variable. These findings suggest that MRC scores should be further investigated to determine their association with LOS. Importantly, our findings have several real-world implications for the identification, clinical management, and potentially prevention of poorer clinical outcomes in critically ill adults with the highest MRC scores.

Firstly, MRC scores can be calculated and incorporated into the EHR to readily identify patients at higher risk. During the COVID-19 pandemic it has become even more evident that the healthcare system and in particular, access and utilization of critical care resources have become profoundly strained[40]. Early identification of higher risk patients based upon MRC scores can aid in triaging limited ICU resources. Currently, there is no standardized MRC method for presenting safety alerts pertaining to medication use. The development and integration of MRC scores into clinical decision support tools can alert interdisciplinary care team members to review and modify the medication regimen to ensure safer, patient-centered care[41–44]. This study corroborates the need of standardizing MRC scores within the critically ill population.

Secondly, our results clearly demonstrate that MRC scores at 24 hours were associated with mortality suggesting they should be incorporated into current practice (**Table 2**). Moreover, our pervious proof-of-concept ML model demonstrated that MRCICU scores are associated with ICU mortality[26]. Moreover, MRC has been shown to be a better predictor of mortality compared to polypharmacy alone[32]. MRC scores at 24 hours were significantly associated with ICU LOS and the incorporation of therapeutic drug classes improved the prediction for LOS and need for MV (**Table 3**). These findings can be confounded however by a) exclusion of the weights for important medications (i.e., vasopressors and anti-infectives) in MRC calculations, b) exclusion of patient’s severity, and c) exclusion of influential combinations of medications regimens.

Thirdly, vasopressors were a significant predictor in all clinical outcome models (**Fig 3**) and were found to be statistically significant in all stepwise models (**Table 2**). In practice, use of vasopressors is indicated in patients with poorer health conditions, such as decompensated heart failure and shock[45,46]. The frequent diagnosis of ICU-related delirium has been a known contributing factor to ICU LOS among other undesirable outcomes[47–49]. Historically, numerous medications have been used to minimize the duration of delirium, yet studies are lacking to identify a safe and effective agent. Our findings of current psychiatric medications potentially contributing to an increase in TOMV suggests the continued need to identify an agent to minimize the incidence and duration of ICU delirium leading to extended TOMV and LOS.

Lastly, the association of pulmonary and paralytic agent use with mortality and the need for MV was anticipated in our findings as these therapeutic classes are commonly associated with high acuity diseases such as acute respiratory distress syndrome (ARDS) and acute brain injury[50,51]. Therefore, our study supports the inclusion of medication class usage to predict critically ill outcomes. Most notably, the lack of medication use into existing severity of illness scoring tools (i.e., SAPS II and APACHE II) are a major shortcoming of their prediction accuracy. MRC scores provide valuable information to bedside clinicians, including critical care pharmacists (CCPs) which have been recognized as essential members of the interdisciplinary care team by major societal organizations[52–54].

Despite finding an association between MRC scores, ICU mortality, LOS, and need for MV in our regression modeling exercise, both scores are not without unique limitations[6,55]. MRCI has been established and validated for use only in the outpatient setting and is not applicable in the critical care setting as it does not incorporate vital critical care medications[16,55]. On the contrary, MRCICU does include critical care medications, yet does not incorporate the complexities of the pharmacotherapeutic regimen such as the medication combinations, dosages, or frequencies. Further, neither previously established scores consider preexisting comorbidities or severity of critical illness, which are crucial when assessing critical care outcomes. Our results support the inclusion of MRC scores into traditional severity index scoring tools (i.e., APACHE II, SAPS II) can improve the prediction of critical care outcomes. We propose adopting MRC scores, preexisting comorbidities, and severity of illness into future modeling to improve the accuracy of prediction.

## Limitations

Our study must be considered in the context of several limitations. First, the retrospective nature of the study design exposes the risk of missing data that can contribute to confounding bias. Second, we were unable to measure the previous exposure of MRC prior to ICU admission due to unconfirmed and variable pre admission medication use. Third, we did not exclude the influence of commonly associated clinical conditions (i.e., ARDS, decompensated heart failure, and sepsis) as contributing factors to MRC. Therefore, the results may be subject to residual and unmeasured confounders. Lastly, both MRC scores at 24 hours and 48 hours can be indirectly related to LOS, as higher LOS may constitute higher MRC scores for the patients who remained admitted to the ICU for more than 48 hours.

## Conclusions

In this retrospective cohort study, our findings suggest that the critical care MRC score should be incorporated into existing severity of illness scoring tools for use in real-time clinical practice. Moreover, incorporation of MRC scores into EHRs and clinical decision support tools will assist all members of the interdisciplinary care team to improve the quality of care through better identification and prediction of higher-risk patients for poorer clinical outcomes.

## Data Availability

The data can be requested from Roger William Medical Medical Center in Rhode Island.

## Supporting information

**S1 Fig.**
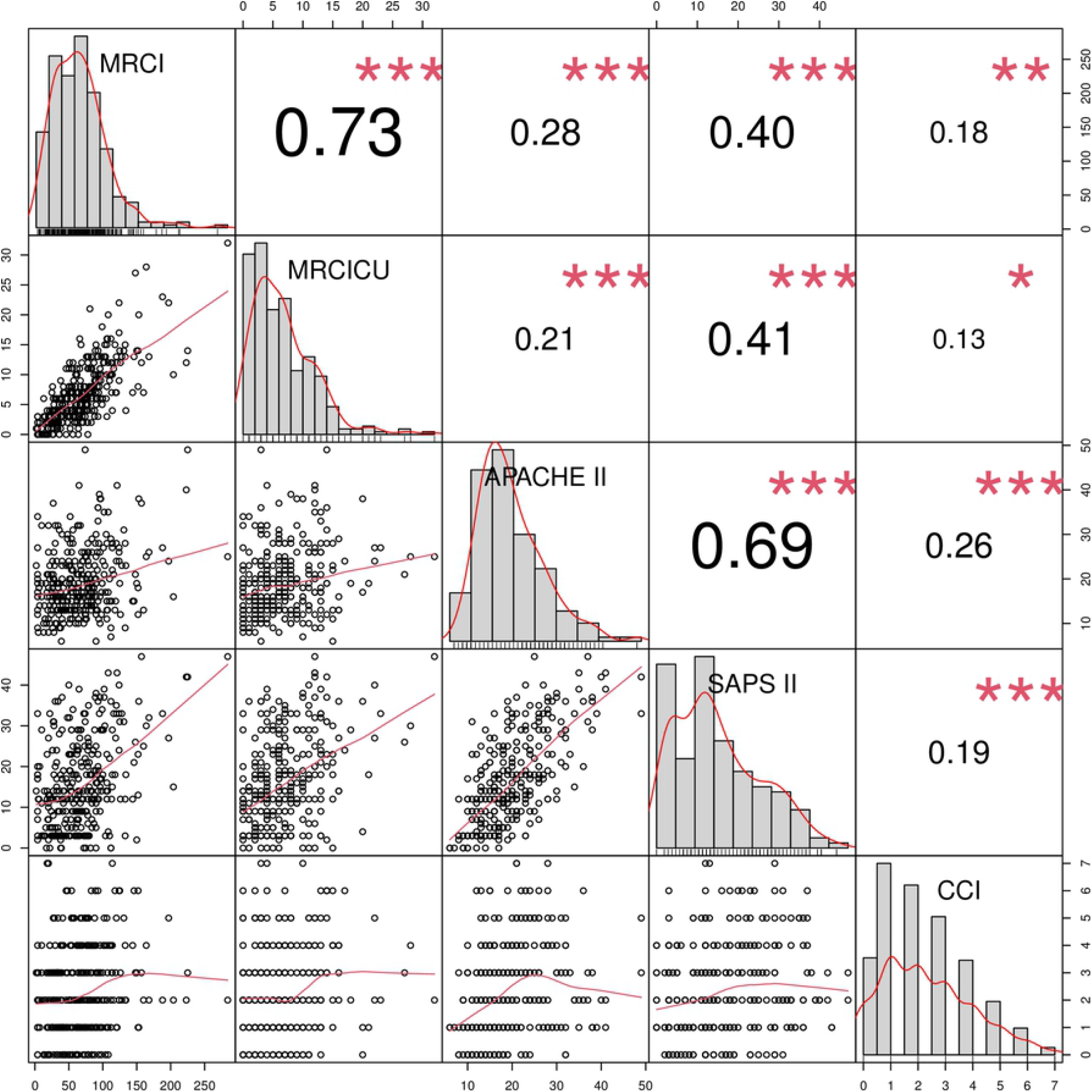
Correlation among the severity and medication regimen scores. Pearson correlation Coefficients for severity of illness scores stratified by age groups: Charlson comorbidity index (CCI), Acute Physiology and Chronic Health Evaluation (APACHE II), Simplified Acute Physiology Score (SAPS) II, Medication Regimen Complexity Index (MRCI), and Medication Regimen Complexity in Intensive Care Unit (MRCICU).

**S2 Fig.**
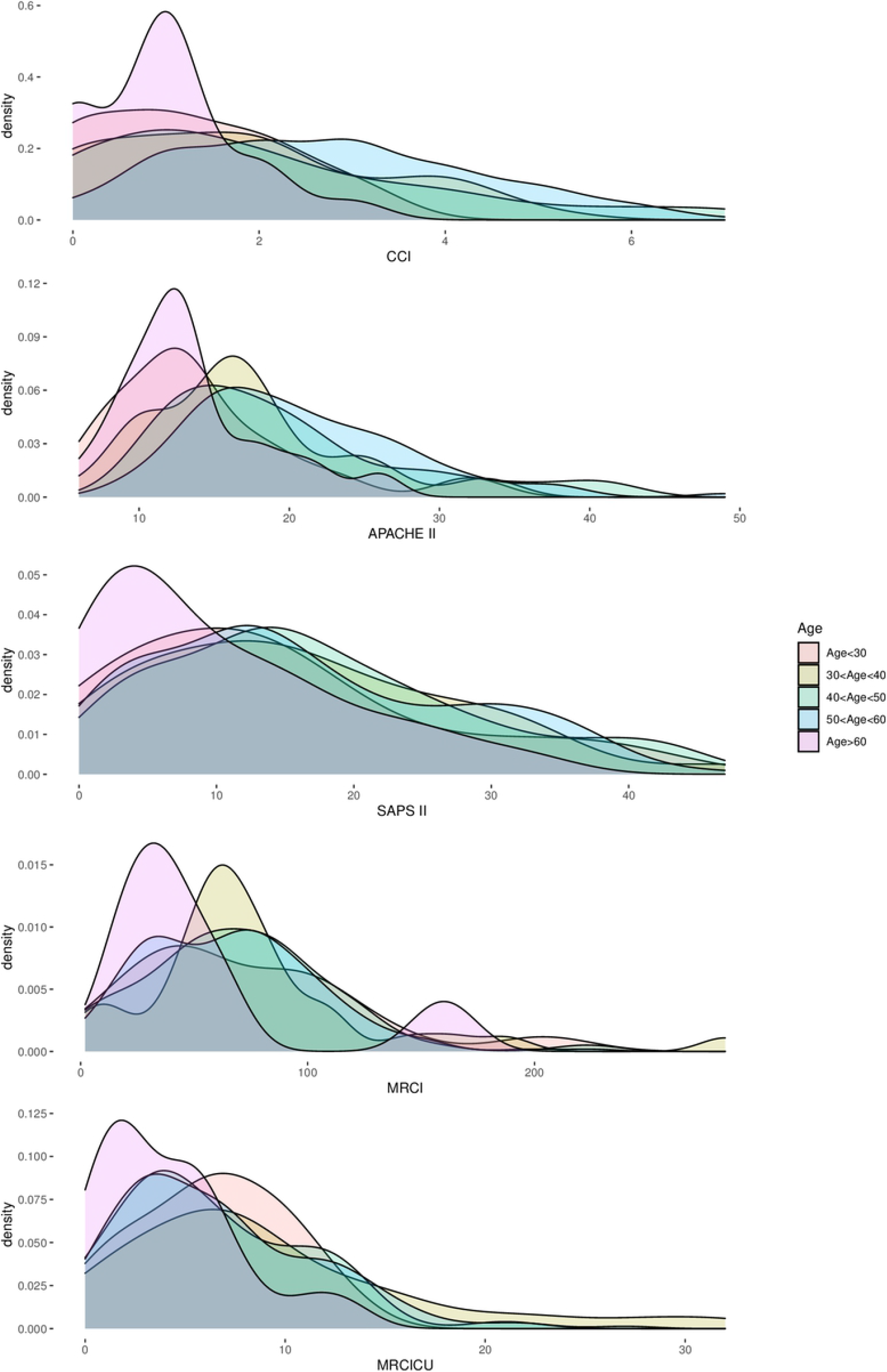
Distribution of the severity and medication regimen scores: Distribution of severity of illness scores stratified by age groups: Charlson comorbidity index (CCI), Acute Physiology and Chronic Health Evaluation (APACHE II), Simplified Acute Physiology Score (SAPS) II, Medication Regimen Complexity Index (MRCI), and Medication Regimen Complexity in Intensive Care Unit (MRCICU).

**S3 Fig.**
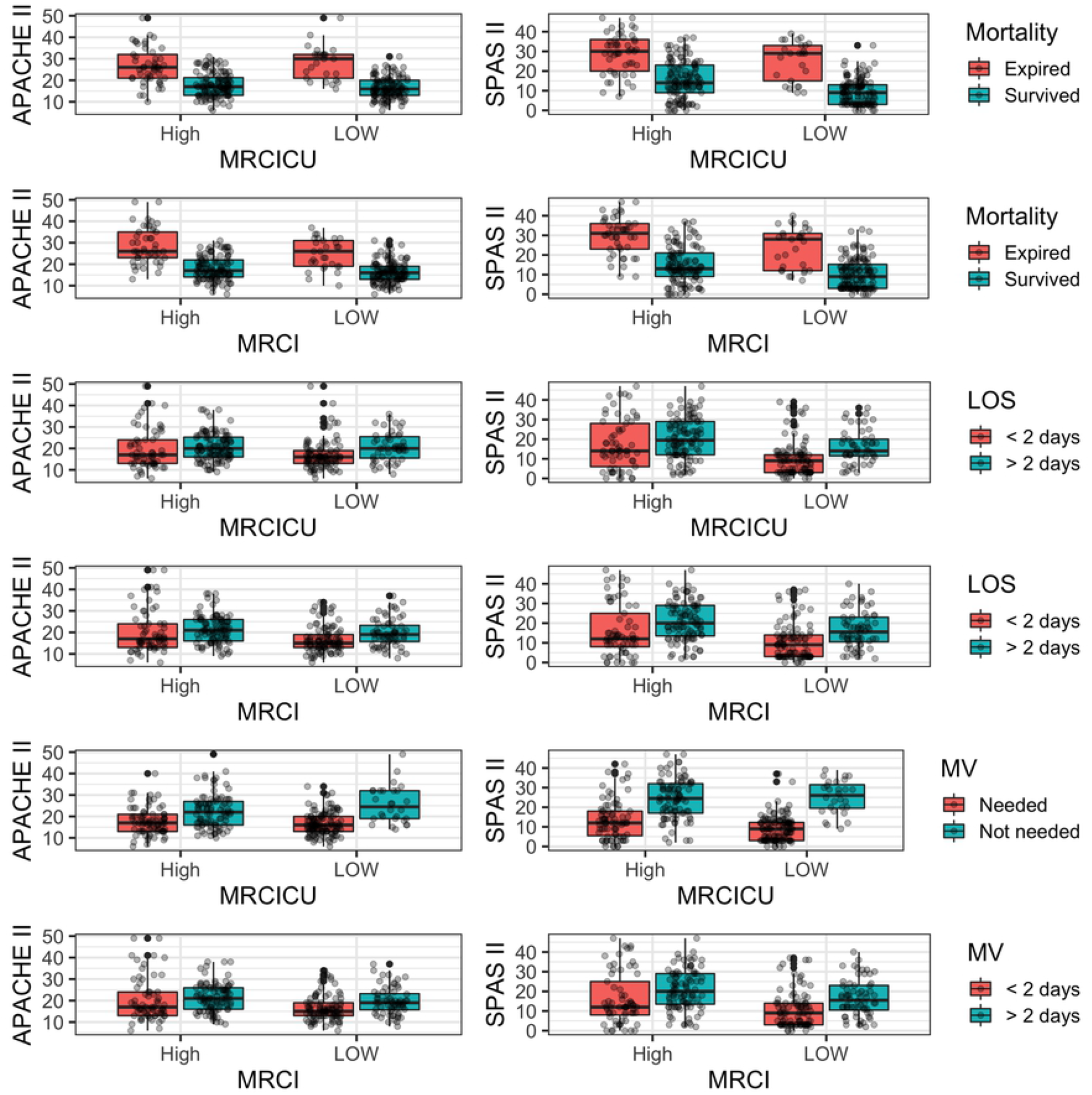
Bivariate analysis of severity of Illness scoring Tools (APACHE II and SAPS II) with MRC Scores (MRCI and MRCICU). Abbreviations: Acute Physiology and Chronic Health Evaluation (APACHE II), Simplified Acute Physiology Score (SAPS) II, Medication Regimen Complexity Index (MRCI), and Medication Regimen Complexity in Intensive Care Unit (MRCICU)

**S1 Table. Variables extracted from electronic health record data**. Description of the variables extracted by group: demographics, vital signs, laboratories, scoring tools, medication classes, and comorbidities.

**S2 Table. Comparison between patient characteristics and Medication Regimen Complexity Index (MRCI) score**. Demographic and clinical characteristics between low and high MRC scoring cohorts.

**S3 Table. Comparison between patient characteristics and Medication Regimen Complexity Intensive Care Unit (MRCICU) score** Demographic and clinical characteristics between low and high MRCICU score cohorts.

**S4 Table. Alternative method for variable selection**. Four L1 penalization techniques (LASSO) for ICU mortality, ICU length of stay, and need for mechanical ventilation.

